# Expression of C5aR1 in Cutaneous Squamous Cell Carcinoma is Associated with Invasion, Metastasis and Poor Prognosis

**DOI:** 10.1101/2024.08.16.24312116

**Authors:** Lauri Heiskanen, Liisa Nissinen, Elina Siljamäki, Jaakko Knuutila, Teijo Pellinen, Markku Kallajoki, Jyrki Heino, Pilvi Riihilä, Veli-Matti Kähäri

**Author notes:** Corresponding author: Prof. Veli-Matti Kähäri, M.D., PhD Department of Dermatology University of Turku and Turku University Hospital Hämeentie 11 TE6 FI-20520 Turku, Finland. **Data availability:** The data underlying this article cannot be shared publicly due to patient data protection regulations. Ethical Issues: The study was approved by The Ethics Committee of the Hospital District of Southwest Finland (187/2006) and Auria Biobank’s Scientific Steering Committee (AB15- 9721). The research was carried out according to Declaration of Helsinki and an informed biobank consent was obtained from the patients. Registry study approval for collection and use of clinical and histopathological data was obtained from the Turku University Hospital Clinical Research Centre (TO5/042/18). All experiments with mice were carried out with permission of the animal test review board of the Southern Finland (ESAVI15107/2020) according to institutional guidelines.

## Abstract

Cutaneous squamous cell carcinoma (cSCC) is the most common metastatic skin cancer, and the metastatic disease is associated with poor prognosis. We have examined the role of complement C5a receptor, C5aR1, in the progression and metastasis of cSCC. C5aR1 expression was increased in cSCC cells in 3D spheroid co-culture model in the presence of fibroblasts, and that treatment with recombinant C5a enhanced the invasion of cSCC cells. Staining for C5aR1 was detected on the surface of tumor cells at the invasive edge of human cSCC xenografts *in vivo*. Staining of metastatic and non-metastatic primary human cSCCs, premalignant and benign epidermal lesions and normal skin for C5aR1 with multiplex immunofluorescence and chromogenic immunohistochemistry revealed increased expression of C5aR1 on the surface of tumor cells and fibroblasts in invasive cSCCs and recessive dystrophic epidermolysis bullosa–associated cSCCs compared to cSCC in situ, actinic keratoses, seborrheic keratoses and normal skin. Increased expression of C5aR1 on the tumor cell surface and in fibroblasts was associated with metastatic risk and poor disease-specific survival of patients with primary cSCC. These findings reveal the role of C5aR1 in cSCC invasion and identify C5aR1 as a novel biomarker for metastasis risk and poor prognosis in patients with cSCC. The results also suggest that C5aR1 could be a novel therapeutic target for treatment of locally advanced and metastatic cSCC.

## Introduction

Cutaneous squamous cell carcinoma (cSCC) is the most common metastatic skin cancer, with an increasing incidence in recent decades.^1–3^ Exposure to solar UV radiation is the predominant risk factor for cSCC, which commonly arises from precursor lesions actinic keratosis (AK) and Bowen’s disease (*in situ* cSCC, cSCCIS) in sun-damaged skin.^4^ Approximately 3-5% of primary cSCCs metastasize and the prognosis for patients with metastatic cSCC (mcSCC) is poor.^5,6^ It is estimated that cSCC accounts for nearly 25% of annual skin cancer deaths.^7^ Currently, there are no established molecular markers in clinical practice for predicting the metastasis risk of primary cSCCs.^7^ Accordingly, there is a need for predictive biomarkers for the prognosis of cSCC and for new therapeutic targets for metastatic cSCC.

Complement system is an integral part of human innate immunity, and its role in cancer progression has recently been emphasized.^8–10^ Tumor cell-specific expression of several complement components (FB, FD, C3, C1r, and C1s) and inhibitors (FH and FI) has been documented in cSCC, and it has been shown that they play a non-canonical role in cSCC progression *in vivo.*^11–17^ Complement component C5a functions as vasodilator and chemotactic factor, and it increases vascular permeability and degranulation of mast cells by attaching to the specific receptor C5aR1 on the target cells.^18–20^ C5aR1 is upregulated in infectious and inflammatory diseases such as sepsis, respiratory distress syndrome, systemic lupus erythematosus and inflammatory bowel disease.^21–23^ Overexpression of C5aR1 has also been shown in several cancer types, including non-small cell lung cancer, urothelial cell carcinoma, renal cell carcinoma, gastric cancer, hepatocellular carcinoma, prostate cancer, and breast cancer^24–32^, and is associated with poor survival.^33^ Additionally, C5a production is upregulated in multiple cancers, and its presence has been linked to increased metastatic potential of cancer cells.^34^ C5a can also modulate the immune microenvironment towards a pro-tumor or anti-tumor response depending on the tumor type and local concentration of C5a.^35^

In our previous studies, the expression of C5aR1 mRNA was not detected in cSCC cells or normal keratinocytes cultured in a monolayer.^12^ The aim of the present study was to further examine the role of C5aR1 in the progression and metastasis of cSCC. The results demonstrate increased expression of C5aR1 in cSCC cells co-cultured with human skin fibroblasts in 3D spheroids and increased invasion of cSCC cells through collagen upon treatment with recombinant C5a. Elevated expression of C5aR1 was noted in the cSCC tumor cells and stromal fibroblasts compared to AK and cSCCIS *in vivo*, with the expression increasing towards mcSCC and cSCC metastases. Moreover, the upregulation of C5aR1 in cSCC cells and fibroblasts in TME was associated with poor prognosis. These findings suggest that C5aR1 could serve as a prognostic biomarker and a therapeutic target for locally advanced and metastatic cSCC.

## Materials and methods

### Cell lines

Primary non-metastatic (UT-SCC-91) and metastatic (UT-SCC-7) cell lines were established from surgically removed cSCCs at Turku University Hospital.^36,37^ The authentication of these cell lines was performed by STR DNA profiling (DDC Medical, Fairfield, OH).^36^ The Ha-*ras*- transformed tumorigenic HaCaT cell line RT3^38^ was kindly provided by Dr. Norbert Fusenig (German Cancer Research Center, Heidelberg, Germany). Primary adult human skin fibroblasts were a kind gift from Prof. Risto Penttinen.^39,40^ Normal human adult dermal fibroblasts (C- 12302) were purchased from PromoCell. Both fibroblast strains were used up to passage number 12. All cell lines were grown in Dulbecco’s modified Eagle’s medium (DMEM with 4.5 g/L glucose; 12-614F, Lonza) supplemented with 10% fetal calf serum (FCS), L-glutamine (6 nmol/L), penicillin (100 U/ml) and streptomycin (100 µg/ml). The cell lines were routinely tested to be negative for mycoplasma contamination using MycoAlert PLUS Mycoplasma Detection Kit.

### Human cSCC xenografts

Human cSCC xenografts were established as previously.^13^ Primary UT-SCC-91 (7 × 10^6^) and metastatic UT-SCC-7 cells (5 × 10^6^) were injected subcutaneously into the back of 6-week-old severe combined immunodeficient (SCID) mice (CB17/Icr-Prkdc^scid^/IcrIcoCrl) (Charles River Laboratories). UT-SCC-91 and UT-SCC-7 xenograft tumors were harvested after 16 or 21 days, respectively, and processed for IHC analysis as described previously.^13^

### Tissue material and chromogenic immunohistochemistry

Tissue samples were collected from Auria Biobank and the archives of University Hospital of Turku. Tissue microarrays (TMAs) consisting of formalin-fixed, paraffin-embedded human tissue specimens obtained by resection or biopsy were constructed, and the characteristics of tumor cohorts were previously described (Table S1).^5,41^ The tissue samples consisted of normal skin (n=54 individual skin samples), seborrheic keratosis (SK; n=6 individual tumors), actinic keratosis (AK; n=50 individual tumors), cSCC in situ (cSCCIS; n=51 individual tumors), non-metastatic cSCC (non-mcSCC; n=152 samples, 97 individual tumors), metastatic cSCC (mcSCC; n=77 samples, 55 individual tumors), cSCC metastases (n=94 samples, 65 individual tumors),and recessive dystrophic epidermolysis bullosa-associated cSCC (RDEBSCC; n=11 individual tumors) were used. TMAs contained replicate cores from each tumor. The findings from tumors with multiple spots were merged to one so that one result per tumor or skin sample was observed.

### Multiplexed immunofluorescence

C5aR1 and activated fibroblast markers, along with their distribution in tumor tissue, were defined using mIF. The first panel contained C5aR1 and fibroblast activation protein (FAP), the second panel CD45 and α-smooth muscle actin (αSMA), and the third staining round PanEpi (Table S2).^41^ C5aR1 expression levels were analyzed based on the relative cell area in normal skin samples (n=62 spots, 54 individual skin samples), actinic keratoses (n=54 spots, 53 individual tumors), cSCCISs (n=47 spots, 47 individual tumors) invasive cSCCs (n=198 spots, 174 individual tumors), cSCC metastases (n=9 spots, 9 individual tumors) and RDEBSCCs (n=83 spots, 11 individual tumors). When multiple spots were derived from the same tumor, only the highest observed result from all the spots was included in the data.

### Statistical Analysis

Statistical analysis was performed as indicated in Supplemental data.

Additional information on Materials and Methods are available in Supplemental data.

## Results

### C5aR1 expression in cSCC cells is induced by co-culture with fibroblasts

Our previous studies revealed that C5aR1 expression is not detected at the mRNA level in cSCC cells cultured in a monolayer.^12^ To further examine the role of C5aR1 in cSCC, we employed co-cultures of cSCC cells and normal human skin fibroblasts in a 3D spheroid model.^39,40^ The results showed that in co-cultures of metastatic cSCC cells (UT-SCC-7) and fibroblasts, C5aR1 expression was markedly increased compared to spheroids that contained only cSCC cells or fibroblasts (Figure 1a, b). Confocal imaging showed peripheral localization of UT-SCC-7 cells, whereas fibroblasts were primarily located in the centre of the spheroid (Figure 1c). Immunofluorescent (IF) staining revealed colocalization of C5aR1-positive cells with UT-SCC-7 cells in the outer shell of the spheroid (Figure 1c). Confocal imaging results were verified by a computational analysis method that allowed the expression profiles of the spheroids to be analyzed, taking into account different diameters and the intensity values of the spheroids. The expression profile analysis showed C5aR1 expression emerging from the spheroid edges, where UT-SCC-7 cells were also located (Figure 1d). To verify the results obtained with UT-SCC-7 cells, Ha-*ras*-transformed metastatic epidermal HaCaT keratinocytes (RT3 cells) were cultured together with human skin fibroblasts. Western blotting showed that C5aR1 expression was increased in spheroids containing both RT3 cells and fibroblasts (Supplementary Figure S1A and S1B). Confocal imaging showed that C5aR1 expression was most prominent in the outer shell of the spheroids, where RT3 cells were also located (Supplementary Figure S1C).

**Figure 1.**
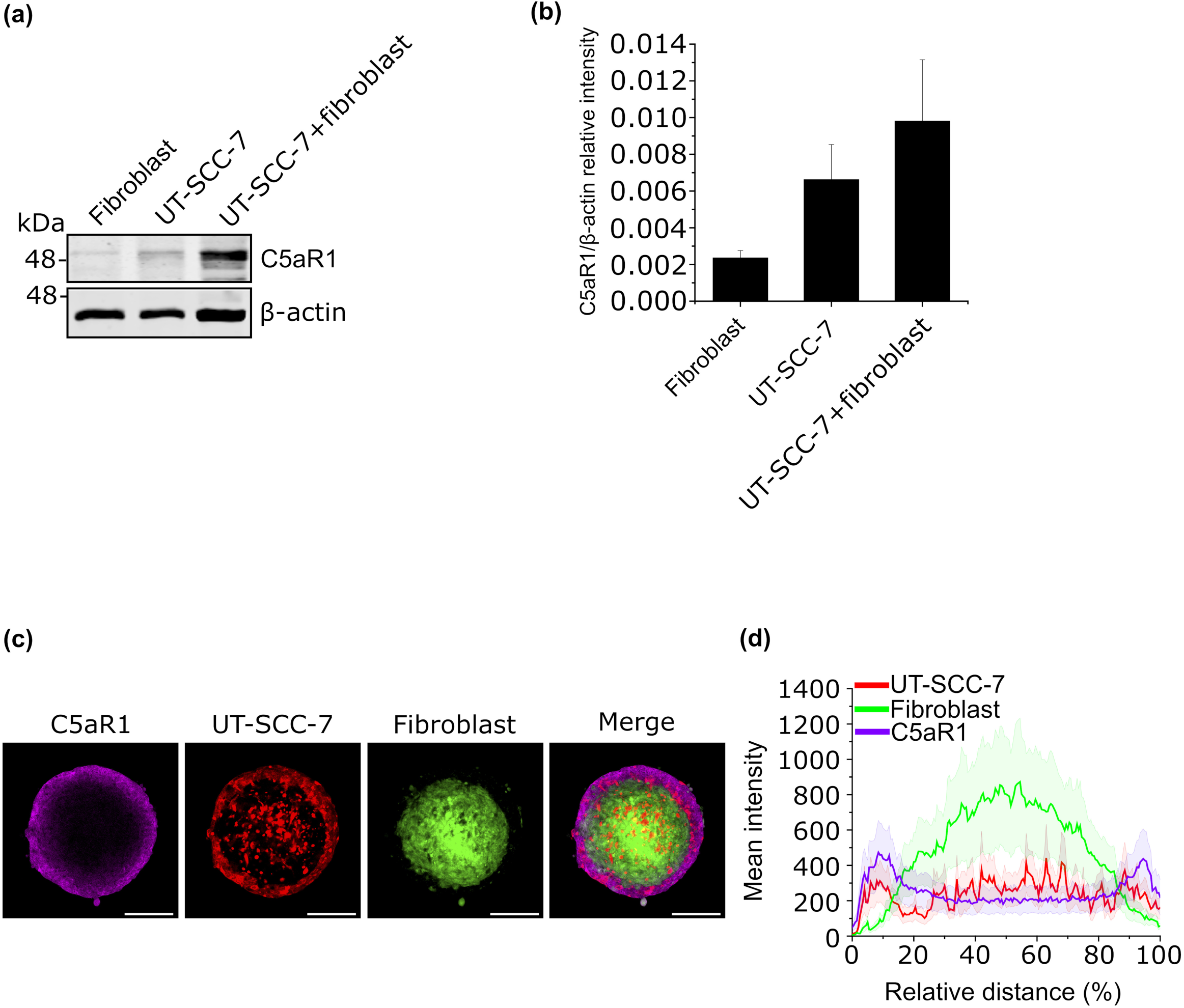
C5aR1 expression in cSCC cells is induced by co-culture with fibroblasts. (a) Western blot analysis of C5aR1 in 3D spheroids composed of human skin fibroblasts, metastastic cSCC cells (UT-SCC-7) and their co-coculture. A representative image from three independent biological replicates is presented. α-actin was used as a loading control. (b) Quantification of C5aR1 levels from the western blots in (a). The graph illustrates C5aR1 relative intensity to β-actin ± SEM. Three independent biological replicates were conducted. (c) Confocal images of spheroids with cSCC cells (UT-SCC-7) co-cultured with human skin fibroblasts. The cells were initially labeled with CellTrackers (UT-SCC-7 cells in red and fibroblasts in green) and the spheroids were allowed to grow for three days. Following PFA fixation, the spheroids were subjected to immunofluorescence staining for C5aR1. Scale bars, 200 µm. Three independent biological replicates were performed, n=24 spheroids. (d) Expression profile of spheroids containing UT-SCC-7 cells and human skin fibroblasts. The cells were treated as in (c), and the expression profile was calculated from the confocal images. Mean intensity values depict the intensities of UT-SCC-7 cells (red), fibroblasts (green) and C5aR1 (magenta), while the relative distance indicates the diameter of the spheroids. 24 spheroids from three biological replicates were analyzed. Mean (dark line) ± SEM (light area around the line) is shown.

### C5a increases RT3 cell invasion in collagen I

To investigate the potential role of C5a in cell invasion, 3D spheroids established with RT3 cells and human skin fibroblasts were treated with recombinant human C5a (rhC5a). Confocal images illustrated that rhC5a treatment enhanced RT3 cell invasion compared to the control cultures (Figure 2a). Quantitative analysis demonstrated that treatment with rhC5a significantly increased the invasion of RT3 cells out of spheroids at the 72-h and 96- h time points compared to untreated control cells (Figure 2b, c). The invasion of fibroblasts in the same spheroids remained unaffected by rhC5a treatment (Figure 2d, e).

**Figure 2.**
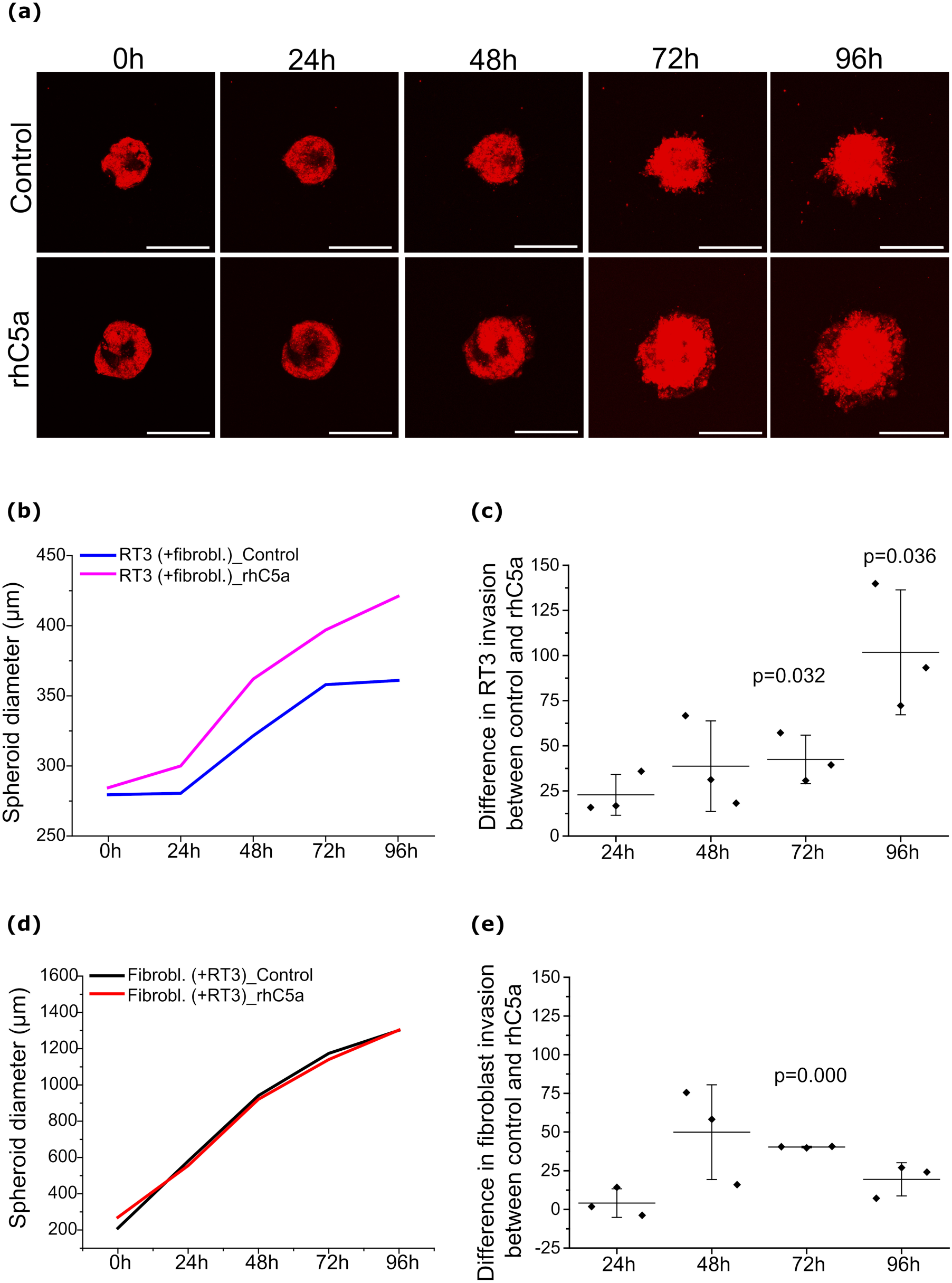
C5a increases RT3 cell invasion in collagen I. (a) RT3 cells (red) were incorporated into spheroids with human skin fibroblasts, and the spheroids were allowed to grow for three days. The spheroids were then treated with recombinant human (rhC5a) for 3 hours, transferred to a 96-well plate, and embedded in a collagen I gel. Invasion was monitored using a confocal microscope every 24 hours for five days. Scale bars, 400 µm. For each time point, 2-6 spheroids were imaged and analyzed. Three independent biological replicates were carried out. (b, c) RT3 cell invasion from co-cultured spheroids that were treated as described in (a). (b) A representative graph from three biological replicates. (c) Analysis of RT3 cell invasion from co-cultured spheroids. The graph illustrates the difference in RT3 cell invasion between control samples and rhC5a- treated samples. The graph displays the mean from three independent biological replicates (squares) ± SD (each replicate contained 2-6 spheroids). p values are from paired t-tests. (d, e) Fibroblast invasion from co-cultured spheroids that were treated as described in (a). (d) A representative graph from three biological replicates. (e) Analysis of fibroblast invasion from co-cultured spheroids. The graph indicates the difference in fibroblast invasion between control samples and rhC5a-treated samples. The graph displays the mean from three independent biological replicates (squares) ± SD (each replicate contained 2-6 spheroids). The p value is from a paired t-test.

### C5aR1 expression on tumor cells in human cSCC xenografts

The expression of C5aR1 *in vivo* was examined using a human cSCC xenograft model. Immunohistochemical (IHC) staining of xenografts showed C5aR1 on the cSCC cell surface, specifically at the edges of xenograft tumors established with both non-metastatic (UT-SCC- 91) (Figure 3a) and metastatic (UT-SCC-7) (Figure 3b) cSCC cell lines. Additionally, in the xenograft tumor established with the metastatic UT-SCC-7 cell line, prominent cell surface staining for C5aR1 was also noted in tumor cells in the center of the tumor (Figure 3b).

**Figure 3.**
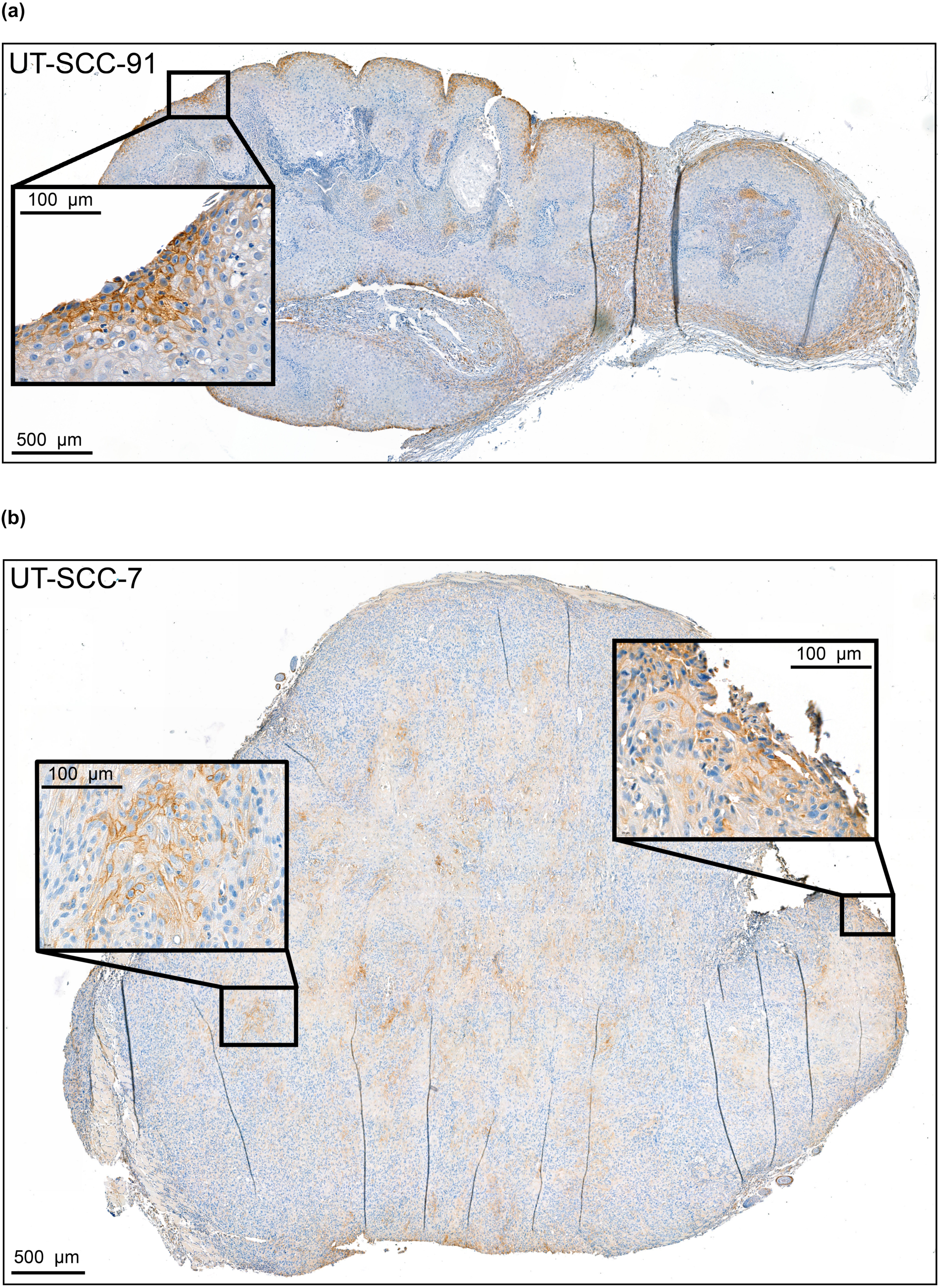
C5aR1 expression on tumour cells in human cSCC xenografts. (a, b) Human non-metastatic (UT-SCC-91, 7x106) and (b) metastatic (UT-SCC-7, 5x106) cSCC cell lines were subcutaneously injected into the backs of severe combined immunodeficient mice. The xenograft tumours were analyzed with immunohistochemistry using C5aR1 antibody. (a) C5aR1 staining in the xenograft tumour established with UT-SCC-91 was predominantly localized at the edges of the xenograft on the tumour cell surface. (b) In the xenograft tumour established with UT-SCC-7, C5aR1 staining was observed on the tumour cell surface at the edges, as well as in the well differentiated areas within the tumour. Cell surface staining appeared more heterogeneous within the tumour, with staining primarily located on the cell surface. Scale bars in the larger panels represent 500 µm, and in smaller panels, 100 µm.

### The increase in C5aR1-positive cells is observed in cSCC *in vivo*

The expression of C5aR1 in invasive cSCCs (n=174) (Figure 4) compared to normal skin (n=62) (Figure 4a), premalignant lesions, actinic keratoses (n=53) (AK, Figure 4b), *in situ* cSCCs (n=47) (cSCCIS, Figure 4c), cSCC metastases (n=9) (Figure 4e) and RDEBSCC (n=11) (Figure 4f) was examined using mIF. The analysis revealed an increased number of C5aR1-positive (C5aR1+) cells in invasive cSCCs compared to normal skin and premalignant lesions (Figure 4a-d). Statistical analysis indicated a significantly higher number of C5aR1+ cells in cSCCs compared to normal skin, AK and cSCCIS (Figure 4g). Multiplex IF for FAP and αSMA, two markers for cancer-associated fibroblasts, demonstrated that the relative abundance of C5aR1+FAP+ cells and C5aR1+SMA+ cells was higher in cSCCs than in normal skin, AK, and cSCCIS (Figure 4a-d, 4h, 4i). Moreover, it was noted that the number of C5aR1+FAP+ and C5aR1+SMA+ cells in cSCC metastases was similar to those in primary cSCC (Figure 4h and i). The assessment of samples of RDEBSCC, an aggressive form of cSCC developing in chronic ulcers of RDEB patients, revealed an elevated number of C5aR1+ cells in RDEBSCC compared to cSCC. Additionally, the number of C5aR1+FAP+ cells was higher in RDEBSCC compared to cSCC (Figure 4h).

**Figure 4.**
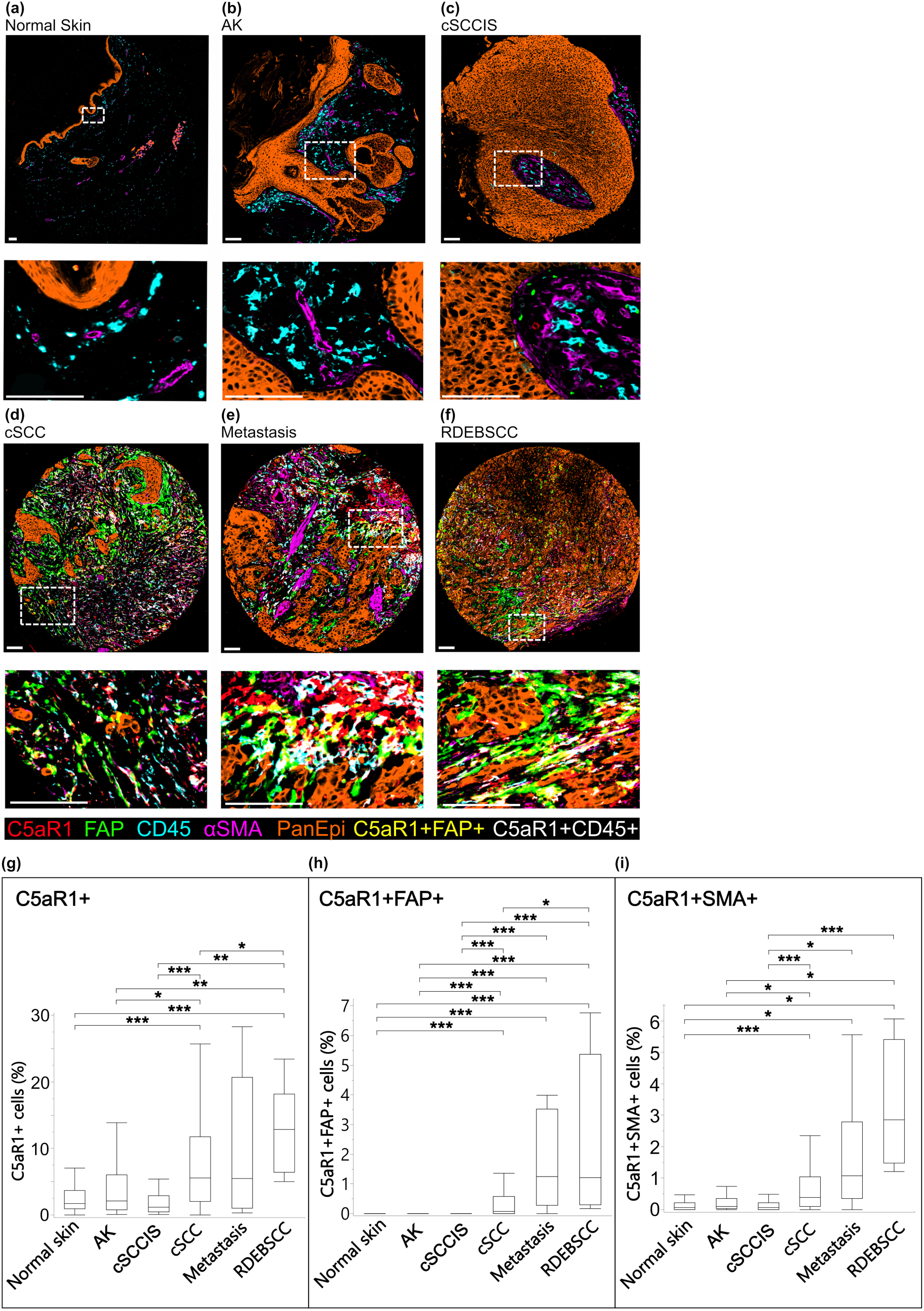
The number of C5aR1-positive cells is increased in cSCC in vivo. (a-f) The expression of C5aR1 in normal skin samples (n=54), actinic keratoses (AK, n=53), cSCC in situ (cSCCISs, n=47), invasive cSCCs (n=174), cSCC metastases (n=9) and recessive dystrophic epidermolysis bullosa associated cSCC (RDEBSCCs, n=11) was analyzed using multiplexed immunofluorescence. Fibroblasts were identified by staining for fibroblast activation protein, (FAP) and α-smooth muscle actin (αSMA). PanEpi staining was used to identify epithelial cells, and CD45 staining to identify leukocytes. Representative images of the stainings from each group are shown. (g) The percentage of C5aR1-positive (C5aR1+) cells in tissue samples. (h) The relative number of C5aR1 and FAP-positive (C5aR1+FAP+) cells in tissue samples. (i) The relative number of C5aR1 and αSMA - positive (C5aR1+SMA+) cells in tissue samples. The figures display the maximum, minimum, upper quartile, lower quartile and median values. The p-values from each pairwise comparison were calculated using the Steel-Dwass method. Significance levels: *p<0.05, **p<0.01, ***p<0.0001.

### C5aR1 is overexpressed on the surface of metastatic cSCC cells and stromal fibroblasts *in vivo*

To further explore the expression and localization of C5aR1, TMAs containing tissue samples of normal skin, SK, AK, cSCCIS, primary non-metastatic and metastatic cSCC, metastasis and RDEBSCC were stained with a C5aR1 antibody using chromogenic IHC. C5aR1 staining was specifically noted on the surface of epithelial cells and with increased staining in cSCC compared to normal skin, SK, AK or cSCCIS (Figure 5). Moreover, increased C5aR1 staining on tumor cell surface was evident in mcSCC (Figure 5b) and cSCC metastases (Figure 5c) compared to non-mcSCC (Figure 5a). Additionally, more abundant C5aR1 staining was observed on tumor cell surface in RDEBSCC (Figure 5d) compared to normal skin (Figure 5e). Interestingly, C5aR1 expression was detected in stromal fibroblasts located near the tumor (Figure 5b, c, d). For semi-quantitative analysis, only fibroblasts in the reticular dermis were included. C5aR1 staining was significantly stronger in TME fibroblasts in non-mcSCCs and mcSCCs compared to normal skin, AKs and cSCCISs samples (Figure 5a, b, e, g, h, j). In normal skin and SK samples all analyzed fibroblasts were negative (Figure 5e, f, j). Additionally, C5aR1 staining on the tumor cell surface and in TME fibroblasts was increased in cSCC metastases compared to non-mcSCC (Figure 5a, b, c, i, j).

**Figure 5.**
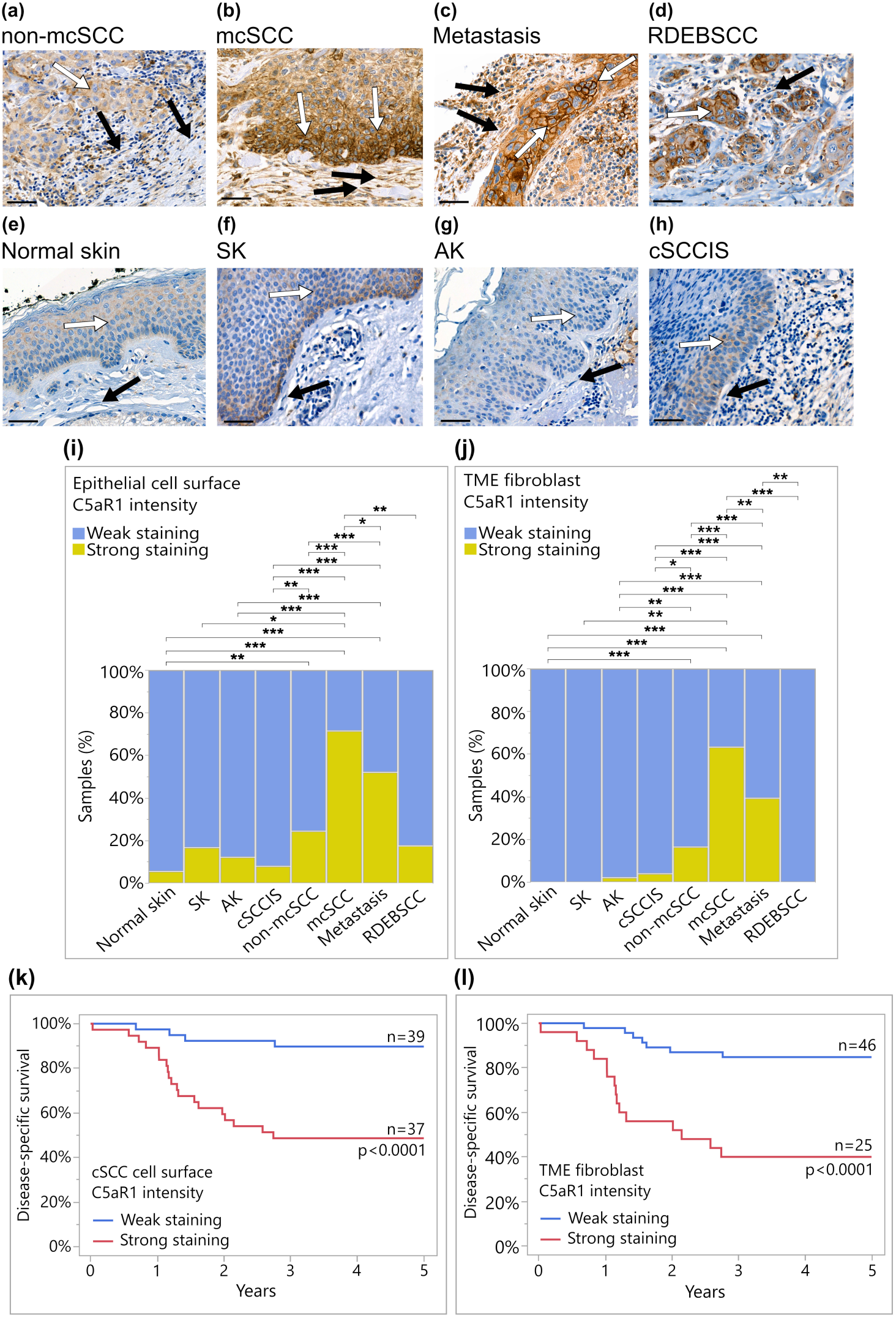
Increased expression of C5aR1 in cSCC in vivo is associated with metastasis and poor prognosis. (a-h) Immunohistochemical staining with a C5aR1 antibody was performed on tissue samples from normal skin (n=54), seborrheic keratosis (SK, n=6), actinic keratosis (AK, n=50), cSCC in situ (cSCCIS, n=51), non-metastatic cSCC (non-mcSCC, n=152 samples, 97 individual tumors), metastatic cSCC (mcSCC, n=77 samples, 55 individual tumors), cSCC metastases (n=94 samples, 65 individual tumors), and recessive dystrophic epidermolysis bullosa-associated cSCC (RDEBSCC, n=11). Representative stainings from each group are shown. Scale bar: 50 µm. Increased C5aR1 staining was observed on the surface of cSCC cell, particularly in mcSCC, metastases and in RDEBSCC (b-d). The C5aR1 staining was weaker on the cell surface in normal skin, SK, AK, cSCCIS and non- mcSCC (a, e-h). In stromal fibroblasts, cytoplasmic C5aR1 staining was strong, especially in mcSCC and metastases (b-c) compared to normal skin, SK, AK, cSCCIS, RDEBSCC and non-mSCC (a,d-h). White arrows indicate tumor and epithelial cells, black arrows indicate fibroblasts in tumor microenvironment (TME). (i, j) C5aR1 immunostaining intensity was scored as weak or strong based on specific staining on the epithelial cell surface and in TME fibroblasts. The C5aR1 staining on the cell surface of cSCC cells (i) and in TME fibroblasts (j) was significantly stronger in non- mcSCC, mcSCC and metastases compared to normal skin, SK, AK and cSCCIS. Additionally, the staining of C5aR1 was stronger on the cSCC cell surface (i) and in fibroblasts (j) in mcSCC and metastases compared to non-mcSCC. Significances: *p<0.05, **p<0.01, ***p<0.0001; Two-tailed Fisher’s Exact Test. (k-l) The Kaplan-Meyer method was utilized to assess cSCC patient survival. (k) Disease-specific survival analysis revealed that increased expression of C5aR1 on the tumor cell surface was associated with poor prognosis in cSCC patients (Log-rank p<0.0001). (l) Strong C5aR1 staining in TME fibroblasts was also associated with poor prognosis in cSCC patients (Log-rank p<0.0001).

### Increased expression of C5aR1 in cSCC *in vivo* is associated with metastasis and poor prognosis

The semi-quantitative analysis of C5aR1 IHC stainings was extended to include survival analysis. The disease-specific 5-year survival analysis revealed that the elevated expression of C5aR1 on the cSCC cell surface (Figure 5k) and in fibroblasts within the TME (Figure 5l), was associated with poor prognosis in cSCC patients. Moreover, this poor prognosis was already evident at 3-years (Figure 5k, l). Furthermore, the impact of C5aR1 on the overall survival of patients with other SCCs was assessed using TCGA database. Elevated expression of *C5AR1* correlated with a poor prognosis, resulting in shorter overall survival in esophageal carcinoma (ESCA) and lung SCC (Supplementary Figure S3).

## Discussion

The complement cascade is part of the human innate immunity system and has conventionally been viewed as a tumor suppressing cytolytic mechanism. However, numerous studies have demonstrated that complement components and their receptors also contribute to tumor progression and metastasis by inducing inflammation or causing immunosuppression.^8–10^ Activation of the complement cascade leads to the cleavage of the complement molecule C5 into C5a and C5b. C5a acts as an anaphylatoxin, whereas C5b is involved in the complement lytic pathway. C5a binds to two receptors C5aR1 and C5aR2 (C5L2) on the surfaces of phagocytes and other cell types.^19,20^ The role of C5aR1 in the inflammatory response is well established, while the function of C5aR2 is less well understood.^20^ Additionally, the role of the C5a-C5aR1 axis in the progression of cSCC is not known.

The aim of this study was to investigate the role of C5aR1 in the progression and metastasis of cSCC. In our previous studies, the expression of C5aR1 at the mRNA level was not detected in cSCC cells cultured in a monolayer.^12^ Here, we observed low expression of C5aR1 at the protein level in a metastatic cSCC cell line (UT-SCC-7) cultured in 3D spheroids. However, when cSCC cells were co-cultured with normal dermal fibroblasts in spheroids, the protein-level expression of C5aR1 in cSCC cells increased. Furthermore, treatment of Ha-*ras*- transformed metastatic human keratinocytes, RT3 cells co-cultured in spheroids with human skin fibroblasts with the C5aR1 ligand rhC5a significantly enhanced the invasion of RT3 cells through collagen I. These results suggest that the interaction between fibroblasts and cancer cells is necessary to induce the expression of C5aR1 by cSCC cells, and that C5aR1 promotes the invasion of cSCC cells. This observation is further supported by the C5aR1 expression noted *in vivo* at the surface of cSCC cells at the invading margin in xenograft tumors established with non-metastatic and metastatic cSCC cell lines. Furthermore, these findings are consistent with previous research on the role of C5aR1 in promoting the invasion of gastric cancer cells.^42^

The expression of C5aR1 was examined in a large panel of tissue samples representing the progression from AKs to invasive and metastatic cSCC, as well as metastases using mIF and chromogenic IHC. The results demonstrated C5aR1 expression on the surface of cSCC tumor cells *in vivo*. C5aR1 expression was significantly higher in cSCC compared to AKs, cSCCISs, normal skin or benign papillomas, SKs. Moreover, C5aR1 expression was increased in mcSCC compared to non-mcSCC and the expression was also significantly higher in metastases than in non-mcSCC. Notably, mIF visualizes the relative cell area positive for C5aR1 and chromogenic IHC the localization and intensity of the staining. These results suggest that C5aR1 could serve as a marker for metastatic primary cSCCs or cSCC metastases. Similar findings have been observed in lung cancer, and meta-analyses have shown a higher level of C5aR1 associated with the occurrence of lymph node metastases.^33,43–45^ Interestingly, in RDEBSCCs the expression of C5aR1 on cSCC tumor cells was comparable to that in non-mcSCC, whereas lesser expression was noted in TME fibroblasts. This suggests that although RDEBSCC is an aggressive form of cSCC there are differences in the expression of C5aR1 positive fibroblasts compared to sporadic cSCCs. These findings are in accordance with our recent observations showing differences in cancer-associated fibroblast (CAF) population between cSCC and RDEBSCC.^41^

The role of CAFs in cancer progression has recently received attention. Our recent observations indicate that FAP and αSMA-positive CAFs are increased in invasive cSCCs.^41^ The findings of this study demonstrate the expression of C5aR1 by TME fibroblasts in cSCC, with an increased co-localization of FAP and αSMA-positive fibroblasts compared to normal skin, AKs and cSCCIS. These results align with previous studies demonstrating that the activation of C3a-C3aR signaling in a mouse breast cancer model resulted in enhanced lung metastasis formation by modulating CAFs.^46^

C5a increased the invasiveness of cSCC cells, and we also observed an upregulation in the expression of C5aR1 on the tumor cell surface as well as in CAFs. These results align with observations in other cancers, such as lung cancer, ^43–45^ and suggest that the C5a-C5aR1 pathway likely contributes to promoting metastasis in cSCC as well. Additionally, our results indicated that high expression of C5aR1 correlated with a poor prognosis. These findings highlight the potential value of C5aR1 expression as a prognostic marker for cSCC, and C5aR1 staining could serve as a predictive biomarker for the prognosis of cSCC patients.

Knocking down the anaphylatoxin-related pathway can result in anti-tumoral effects in many cancers.^34^ The potential mechanisms that may underlie the correlation between the metastatic potential of cSCC and the overexpression of C5aR1 could be linked to the established functions of anaphylatoxins. Anaphylatoxins are vasodilators and increase the vascular permeability.^18^ This function of anaphylatoxins could enhance the metastatic potential of cSCC tumors by increasing the permeability of capillaries in the TME. Anaphylatoxins can also serve as promoters of chronic inflammation in the TME, thereby promoting tumor progression.^8–11^

The C5a/C5aR1 signaling pathway plays an important role in the inflammatory cell signaling within the TME, and this pathway has been recognized as a potential therapeutic target in the context of checkpoint inhibition^42,44^. It has been shown that inhibiting C5a/C5aR signaling improves the efficacy of PD-1 blockade, resulting in a significant reduction in tumor growth and metastasis, along with prolonged survival in lung cancer patients.^43^ This finding is particularly intriguing in the context of treating advanced cSCC.

Currently, there are only two immune-oncological PD-1 antibody treatments available for locally advanced and metastatic cSCCs targeting the PD-1/PDL-1 pathway: cemiplimab, which is approved by the FDA and EMA, and pembrolizumab, which is FDA-approved.^47–49^ The findings of our study suggest that the C5a-C5aR1 axis could potentially serve as a therapeutic target in cSCC in combimation with PD-1 antibody treatment. Currently, three antibodies against C5a and one small molecule inhibitor against C5aR1 have been approved for treatment of inflammatory diseases.^50^ It is conceivable that these therapies could also represent viable options for treating solid cancers including advanced cSCC.

In conclusion, our results demonstrate elevated C5aR1 expression in cSCC tumors, particularly at the invasive tumor edges and in stromal fibroblasts, compared to normal skin, benign papillomas, AKs or cSCCISs. C5a promotes cSCC cell invasion, and the expression of C5aR1 is linked to metastatic risk and poor prognosis in cSCC patients. These findings suggest that C5aR1 could serve as a potential metastatic risk marker, a novel prognostic biomarker, and promising therapeutic target for cSCC.

## Disclosure statement

The authors state no conflict of interest

## Supplemental data

Supplemental material for this article can be found at http://doi.org/

## Data Availability

The data underlying this article cannot be shared publicly due to patient data protection regulations.

## Acknowledgements

We thank Johanna Markola for skillful technical assistance. The authors thank Annabrita Schoonenberg (FIMM) for performing the mIF stainings and the staff at the Histology Core of the Institute of Biomedicine at the University of Turku. ChatGPT-4 was used to revise the language of some paragraphs during the final editing the manuscript.

---

**Figure 6.**
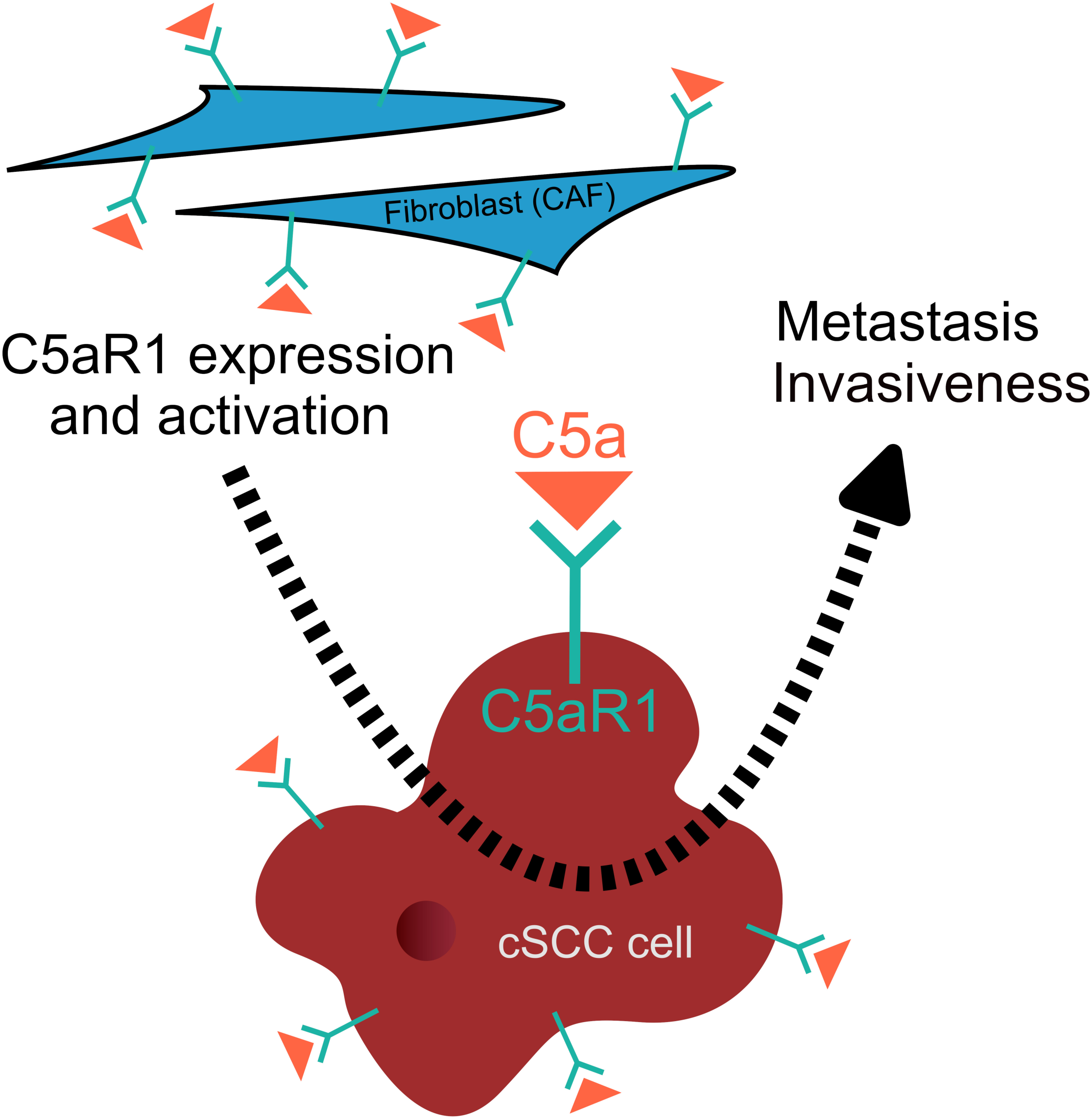
Graphical Abstract. Our study demonstrates that the overexpression of C5aR1 in cSCC cells is associated with increased invasiveness and metastatic potential in cSCC. Additionally, fibroblasts in the tumour microenvironment (TME) show increased C5aR1 expression, which also contributes to the invasiveness of cSCC.

## Notes

**Funding sources:** The study was supported by Sigrid Jusélius Foundation, Jane and Aatos Erkko Foundation, Finnish Cancer Foundation, Cancer Foundation of the Southwest Finland, Finnish Dermatological Society, The Maud Kuistila Memorial Foundation, Ida Montin Foundation, The Finnish Medical Foundation, Finnish Cultural Foundation, Päivikki and Sakari Sohlberg Foundation and Turku University Hospital VTR grants (project number: 13336). LH is doctoral candidate in the Doctoral Program for Clinical Investigation of the University of Turku

### Competing Interest Statement

The authors have declared no competing interest.

### Funding Statement

The study was supported by Sigrid Juselius Foundation, Jane and Aatos Erkko Foundation, Finnish Cancer Foundation, Cancer Foundation of the Southwest Finland, Finnish Dermatological Society, The Maud Kuistila Memorial Foundation, Ida Montin Foundation, The Finnish Medical Foundation, Finnish Cultural Foundation, Paivikki and Sakari Sohlberg Foundation and Turku University Hospital VTR grants (project number: 13336).

### Author Declarations

The study was approved by The Ethics Committee of the Hospital District of Southwest Finland and Auria Biobank Scientific Steering Committee.

## References

1. Nehal KS, Bichakjian CK. Update on keratinocyte carcinomas. N Engl J Med 2018; 379:363– 74.

2. Venables ZC, Autier P, Nijsten T, et al. Nationwide incidence of metastatic cutaneous squamous cell carcinoma in England. JAMA Dermatol 2019; 155:298.

3. Venables ZC, Nijsten T, Wong KF, et al. Epidemiology of basal and cutaneous squamous cell carcinoma in the U.K. 2013–15: a cohort study. Br J Dermatol 2019; 181:474–82.

4. Ratushny V, Gober MD, Hick R, et al. From keratinocyte to cancer: the pathogenesis and modeling of cutaneous squamous cell carcinoma. J Clin Invest 2012; 122:464–72.

5. Knuutila J, Riihilä P, Kurki S, et al. Risk factors and prognosis for metastatic cutaneous squamous cell carcinoma: A cohort study. Acta Derm Venereol 2020; 100:1–9.

6. Nagarajan P, Asgari MM, Green AC, et al. Keratinocyte carcinomas: Current concepts and future research priorities. Clin Cancer Res 2019; 25:2379–91.

7. Stratigos AJ, Garbe C, Dessinioti C, et al.; EADO, EDF, ESTRO, UEMS, EADV and EORTC. European consensus-based interdisciplinary guideline for invasive cutaneous squamous cell carcinoma. Part 1: Diagnostics and prevention-Update 2023. Eur J Cancer 2023(I); 193:113251

8. Pio R, Corrales L, Lambris JD. The role of complement in tumor growth. Adv Exp Med Biol 2014; 772:229.

9. Riihilä P, Nissinen L, Knuutila J, et al. Complement system in cutaneous squamous cell carcinoma. Int J Mol Sci 2019; 20:3550.

10. Meri S, Magrini E, Mantovani A, Garlanda C. The yin yang of complement and cancer. Cancer Immunol Res 2023; 11:1578–88.

11. Riihilä PM, Nissinen LM, Ala-aho R, et al. Complement factor H: a biomarker for progression of cutaneous squamous cell carcinoma. J Invest Dermatol 2014; 134:498–506.

12. Riihilä P, Nissinen L, Farshchian M, et al. Complement factor I promotes progression of cutaneous squamous cell carcinoma. J Invest Dermatol 2015; 135:579–88.

13. Riihilä P, Nissinen L, Farshchian M, et al. Complement component C3 and complement factor B promote growth of cutaneous squamous cell carcinoma. Am J Pathol 2017; 187:1186–1197

14. Riihilä P, Viiklepp K, Nissinen L, et al. Tumor-cell-derived complement components C1r and C1s promote growth of cutaneous squamous cell carcinoma. Brit J Dermatol 2020; 182:658–70.

15. Rahmati Nezhad P, Riihilä P, Piipponen M, et al. Complement factor I upregulates expression of matrix metalloproteinase-13 and -2 and promotes invasion of cutaneous squamous carcinoma cells. Exp Dermatol 2021; 30:1631–1641

16. Viiklepp K, Nissinen L, Ojalill M, et al. C1r upregulates production of matrix metalloproteinase-13 and promotes invasion of cutaneous squamous cell carcinoma. J Invest Dermatol 2022; 142:1478–1488.e9

17. Rahmati Nezhad P, Riihilä P, Knuutila JS, et al.. Complement factor D is a novel biomarker and putative therapeutic target in cutaneous squamous cell carcinoma. Cancers 2022; 14:305

18. Peng Q, Li K, Sacks SH, Zhou W. The role of anaphylatoxins C3a and C5a in regulating innate and adaptive immune responses. Inflamm Allergy Drug Targets 2009; 8:236–46.

19. Lee H, Whitfeld PL, Mackay CR. Receptors for complement C5a. The importance of C5aR and the enigmatic role of C5L2. Immunol Cell Biol 2008; 86:153–60.

20. Ward PA. Functions of C5a receptors. J Mol Med (Berl) 2009; 87:375–8.

21. Huber-Lang MS, Younkin EM, Sarma JV, et al. Complement-induced impairment of innate immunity during sepsis. J Immunol 2002; 169:3223–31.

22. Sarma VJ, Huber-Lang M, Ward PA. Complement in lung disease. Autoimmunity 2006; 39:387–94.

23. Hopkins P, Michael Belmont H, Buyon J, et al. Increased levels of plasma anaphylatoxins in systemic lupus erythematosus predict flares of the disease and may elicit vascular injury in lupus cerebritis. Arthritis Rheum 1988; 31:632–41.

24. Gu J, Ding J yong, Lu C lai, et al. Overexpression of CD88 predicts poor prognosis in non-small-cell lung cancer. Lung Cancer 2013; 81:259–65.

25. Wada Y, Maeda Y, Kubo T, et al. C5a receptor expression is associated with poor prognosis in urothelial cell carcinoma patients treated with radical cystectomy or nephroureterectomy. Oncol Lett 2016; 12:3995.

26. Maeda Y, Kawano Y, Wada Y, et al. C5aR is frequently expressed in metastatic renal cell carcinoma and plays a crucial role in cell invasion via the ERK and PI3 kinase pathways. Oncol Rep 2015; 33:1844–50.

27. Xi W, Liu L, Wang J, et al. Enrichment of C5a-C5aR axis predicts poor postoperative prognosis of patients with clear cell renal cell carcinoma. Oncotarget 2016; 7:80925.

28. Kaida T, Nitta H, Kitano Y, et al. C5a receptor (CD88) promotes motility and invasiveness of gastric cancer by activating RhoA. Oncotarget 2016; 7:84798.

29. Nitta H, Shimose T, Emi Y, et al. Expression of the anaphylatoxin C5a receptor in gastric cancer: implications for vascular invasion and patient outcomes. Medical Oncol 2016; 33:1–10.

30. Hu WH, Hu Z, Shen X, et al. C5a receptor enhances hepatocellular carcinoma cell invasiveness via activating ERK1/2-mediated epithelial–mesenchymal transition. Exp Mol Pathol 2016; 100:101–8.

31. Imamura R, Kitagawa S, Kubo T, et al. Prostate cancer C5a receptor expression and augmentation of cancer cell proliferation, invasion, and PD-L1 expression by C5a. Prostate 2021; 81:147–56.

32. Imamura T, Yamamoto-Ibusuki M, Sueta A, et al. Influence of the C5a–C5a receptor system on breast cancer progression and patient prognosis. Breast Cancer 2016; 23:876–85.

33. Wang Z, Yu W, Qiang Y, et al. Clinicopathological features and prognostic significance of C5aR in human solid tumors: a Meta-analysis. BMC Cancer 2021; 21:1136.

34. Ajona D, Ortiz-Espinosa S, Pio R. Complement anaphylatoxins C3a and C5a: Emerging roles in cancer progression and treatment. Semin Cell Dev Biol 2019; 85:153–63.

35. Roumenina, Daugan, Petitprez, et al. Context-dependent roles of complement in cancer. Nat Rev Cancer 2019; 19:698–715.

36. Farshchian M, Nissinen L, Grénman R, Kähäri VM (2017) Dasatinib promotes apoptosis of cutaneous squamous carcinoma cells by regulating activation of ERK1/2 2017; Exp Dermatol 26:89–92

37. Nissinen L, Riihilä P, Viiklepp K, Rajagopal V, Storek M, Kähäri VM. (2024) C1s targeting antibodies inhibit the growth of cutaneous squamous carcinoma cells. Sci Rep 2024; 14:13465

38. Boukamp P, Stanbridge EJ, Foo DY, et al. c-Ha-ras oncogene expression in immortalized human keratinocytes (HaCaT) alters growth potential in vivo but lacks correlation with malignancy. Cancer Res 1990; 50:2840–7.

39. Siljamäki E, Rappu P, Riihilä P, et al. H-Ras activation and fibroblast-induced TGF-β signaling promote laminin-332 accumulation and invasion in cutaneous squamous cell carcinoma. Matrix Biol 2020; 87:26–47.

40. Siljamäki E, Riihilä P, Suwal U, et al. Inhibition of TGF-β signaling, invasion and growth of cutaneous squamous cell carcinoma by PLX8394. Oncogene 2023; 42:3633–3647

41. Knuutila JS, Riihilä P, Nissinen L, et al. Cancer-associated fibroblast activation predicts progression, metastasis and prognosis of cutaneous squamous cell carcinoma. Int J Cancer 2024; Apr 22. doi: 10.1002/ijc.34957. Online ahead of print.

42. Nitta H, Wada Y, Kawano Y, et al. Enhancement of human cancer cell motility and invasiveness by anaphylatoxin C5a via aberrantly expressed C5a receptor (CD88). Clin Cancer Res 2013; 19:2004–13.

43. Ajona D, Ortiz-Espinosa S, Moreno H, et al. A combined PD-1/C5a blockade synergistically protects against lung cancer growth and metastasis. Cancer Discov 2017; 7:694–703.

44. Ajona D, Zandueta C, Corrales L, et al. Blockade of the complement C5a/C5aR1 axis impairs lung cancer bone metastasis by CXCL16-mediated effects. Am J Respir Crit Care Med 2018; 197:1164–76.

45. Vadrevu SK, Chintala NK, Sharma SK, et al. Complement c5a receptor facilitates cancer metastasis by altering T-cell responses in the metastatic niche. Cancer Res 2014; 74:3454–65

46. Shu C, Zha H, Long H, et al. C3a-C3aR signaling promotes breast cancer lung metastasis via modulating carcinoma associated fibroblasts. J Exp Clin Cancer Res 2020; 39:11.

47. Migden MR, Rischin D, Schmults CD, et al. PD-1 Blockade with Cemiplimab in Advanced Cutaneous Squamous-Cell Carcinoma. N Engl J Med 2018; 379:341–51.

48. Peris K, Piccerillo A, Del Regno L, Di Stefani A. Treatment approaches of advanced cutaneous squamous cell carcinoma. J Eur Acad Dermatol Venereol 2022; 36 Suppl 1:19–22.

49. Stratigos AJ, Garbe C, Dessinioti C, et al. EADO, EDF, ESTRO, UEMS, EADV and EORTC. European consensus-based interdisciplinary guideline for invasive cutaneous squamous cell carcinoma: Part 2. Treatment-Update 2023(II). Eur J Cancer 2023; 193:113252.

50. West EE, Woodruff T, Fremeaux-Bacchi V, Kemper C. Complement in human disease: approved and up-and-coming therapeutics. Lancet 2024; 403:392–405.

