## Supplemental data for "Expression of C5aR1 in Cutaneous Squamous Cell Carcinoma is Associated with Invasion, Metastasis and Poor Prognosis"

[1]Department of Dermatology, University of Turku and Turku University Hospital, Turku, Finland; [2]FICAN West Cancer Centre Research Laboratory, University of Turku and Turku University Hospital, Turku, Finland. [3]Department of Life Technologies and InFLAMES Research Flagship, University of Turku, Turku, Finland; [4]MediCity Research Laboratory University of Turku, Turku, Finland; [5]Institute for Molecular Medicine Finland (FIMM), Helsinki Institute of Life Science (HiLIFE), University of Helsinki, Helsinki, Finland; [6]Department of Pathology, University of Turku and Turku University Hospital, Turku, Finland.

#### Content:

#### Supplemental Materials and methods

#### Supplemental Figures and Tables

**Figure S1.** C5aR1 expression in RT3 cells is induced by co-culture with fibroblasts.
**Figure S2.** C5aR1 staining in epithelial cell surface and in TME fibroblasts.
**Figure S3.** Correlation of the expression of *C5AR1* with a prognosis of the patients with esophageal carcinoma and lung SCC.
**Table S1.** Baseline characteristics of patients and tumors.
**Table S2.** Antibodies used in this study.

### Supplemental Materials and methods

#### 3D spheroid cultures

The information below on experimental parameters, spheroid preparation and growth conditions is based on MISpheroID guidelines and recommendations.[1] For western blots and invasion assays, 3D spheroids were made in micro-molds according to the manufacturer's instructions (MicroTissues 3D Petri Dish micro-mold spheroids, Sigma-Aldrich) with $2.5 \times 10^5$ cells in one mold (monocultures; 7000 cells/spheroid) or $5.0 \times 10^5$ cells in one mold (cocultures; 14 000 cells/spheroid). In cocultures the cell ratio was 1:1 (RT3/UT-SCC-7 cells and fibroblasts, respectively). The spheroids were grown in serum-free DMEM medium for three days at 37 °C in an incubator environment of 20% $O_2$ and 5% $CO_2$. Ascorbic acid (50 µg/ml) was added daily.

#### CellTracker labeling

RT3 cells, UT-SCC-7 cells and human skin primary fibroblasts were labeled with CellTrackers in 2D condition; RT3 cells and UT-SCC-7 in red (CellTracker Orange CMTMR Dye, C2927, Invitrogen) and fibroblasts in green (CellTracker Green CMFDA Dye, C2925, Invitrogen). The cells were labeled with 2.5 µM dye in DMEM supplemented with 10% FCS for 1 h /+37°C, washed twice with PBS and constructed into spheroids. After three days, the spheroids were used in invasion assays or fixed for immunofluorescence staining.

#### Immunofluorescence of spheroids

Three-day-old spheroids were fixed with 4% PFA for 1h/+4°C, followed by fixation with 4% PFA supplemented with 1% Triton X-100 for 1 h/+4°C. The spheroids were washed twice with PBST (PBS + 0.1% Triton X-100) and blocked with 6% bovine serum albumin in PBST for 3h at room temperature (RT; 20 - 25 °C). C5aR1 antibody (ab234757, Abcam; 1:100 in PBST) was incubated for 3h at RT, and spheroids were then washed three times with PBST (15 min each wash). After that the spheroids were treated with highly pre-cross absorbed Alexa Fluor 633 goat anti-rabbit IgG secondary antibody (A21071, Invitrogen; 1:200 in PBST) for 1.5 h at RT and washed three times with PBST (15 min each wash). The spheroids were mounted in 95% glycerol.

#### Invasion assays from 3D spheroids in collagen I

The cells were stained with CellTrackers as described above, constructed into spheroids and allowed to grow for three days. Ascorbic acid (50 µg/ml in serum-free DMEM medium) was added daily. Three-day-old spheroids were plated on collagen I coated 96-well plates (0.035 mg/ml; Collagen solution from bovine skin, C4243, Sigma) and collagen I gel (2.0 mg/ml; Type I Bovine Collagen Solution, 5010, Nutragen) was layered on the top of the spheroids. DMEM supplemented with 10% FCS was added above the collagen gel. To study the effect of C5a on cell invasion, three-days old spheroids were incubated with 100 nM recombinant human complement component C5a protein (#2037-C5, R&D Systems) for 3 h at 37°C before embedding the spheroids into the collagen gel. Control samples were incubated with 0.1% BSA-PBS for 3 h at 37°C. The collagen I gel and the medium on top of the collagen I gel were supplemented with 100 nM rhC5a, and in control samples with 0.1% BSA-PBS. Spheroids were allowed to invade for 96 h and they were imaged every 24 h with Zeiss LSM880 confocal microscope. Fiji, an image processing platform based on ImageJ [2] was used to calculate the invasion, *i.e.* the area covered by cells. The cell invasion of 2-6 spheroids from each sample were analysed from three independent biological replicates.

#### Confocal imaging

The spheroids were imaged with Zeiss LSM880 AiryScan confocal microscope (Zeiss, Jena, Germany) (10x and 20x objectives; green CellTracker excitation at 488 nm, orange CellTracker excitation at 543 nm, and Alexa Fluor 633-secondary antibody excitation at 633 nm). The imaging was performed at the Cell Imaging and Cytometry Core, Turku Bioscience Centre, Turku, Finland, with the support of Biocenter Finland.

#### Gene expression profiling interactive analysis

The online Gene Expression Profiling Interactive Analysis (GEPIA; http://gepia.cancer-pku.cn/) analysis tool was utilized to analyze the relationship between *C5AR1* expression and prognosis of esophageal carcinoma (ESCA) and lung SCC (LUSC) in The Cancer Genome Atlas (TCGA) data[3,4].

#### Statistical analysis

All quantitative data regarding coculture models are presented as mean ± standard deviation (SD) or standard error of the mean (SEM) as stated in the figure legends. Shapiro Wilk test was used to test normality assumption. Levene's test was used to test the homogeneity of variances between the statistically compared groups. Statistical differences were determined using either paired *t*-test or analysis of variance (ANOVA) complemented by appropriate post hoc tests (Tukey (if variances between the statistically compared groups were similar) or Dunnett's T3 (if variances between statistically compared groups were not similar)). Origin 2015 software (OriginLab Corporation) and SPSS (IBM Corporation, version 25) were used to perform the analyses. Only two-tailed p values $< 0.05$ were considered as statistically significant.

Statistical analysis of immunohistochemical staining and survival analysis were made with JMP®, Version Pro 16. SAS Institute Inc., Cary, NC, 1989–2023. The semiquantitative analysis included four classes based on C5aR1 staining rate; "-" for negative staining, "+" for weak staining, "++" for moderate staining and "+++" for strong staining. For statistical analysis these classes were grouped in two classes, as "weak staining" including "-" and "+" groups and "strong staining" including "++" and "+++" groups. The statistical analysis of semiquantitative data was made with Fischer's Exact test.  Survival analysis was made with Kaplan-Meyer curves and Log-rank analysis. Only two-tailed p-values $< 0.05$ were considered as statistically significant. For multiplexed immunofluorescence the statistical analyses were made with Kruskal-Wallis -test and comparisons with for each pair was implemented with Steel-Dwass method.

#### Supplemental references

1. Peirsman A, Blondeel E, Ahmed T, *et al*. MISpheroID: a knowledgebase and transparency tool for minimum information in spheroid identity. *Nature Methods* 2021 **18**:11 2021; 18:1294–303.
2. Schindelin J, Arganda-Carreras I, Frise E, *et al.* Fiji: an open-source platform for biological-image analysis. *Nat Methods* 2012; **9**:676–82.
3. Uhlen M, Zhang C, Lee S, *et al.* A pathology atlas of the human cancer transcriptome. *Science* 2017; 357:eaan2507.
4. Cancer Genome Atlas Research Network, Weinstein JN, Collisson EA, *et al.* The Cancer Genome Atlas pan-cancer analysis project. *Nat Genet*. 2013; 45:1113-20.

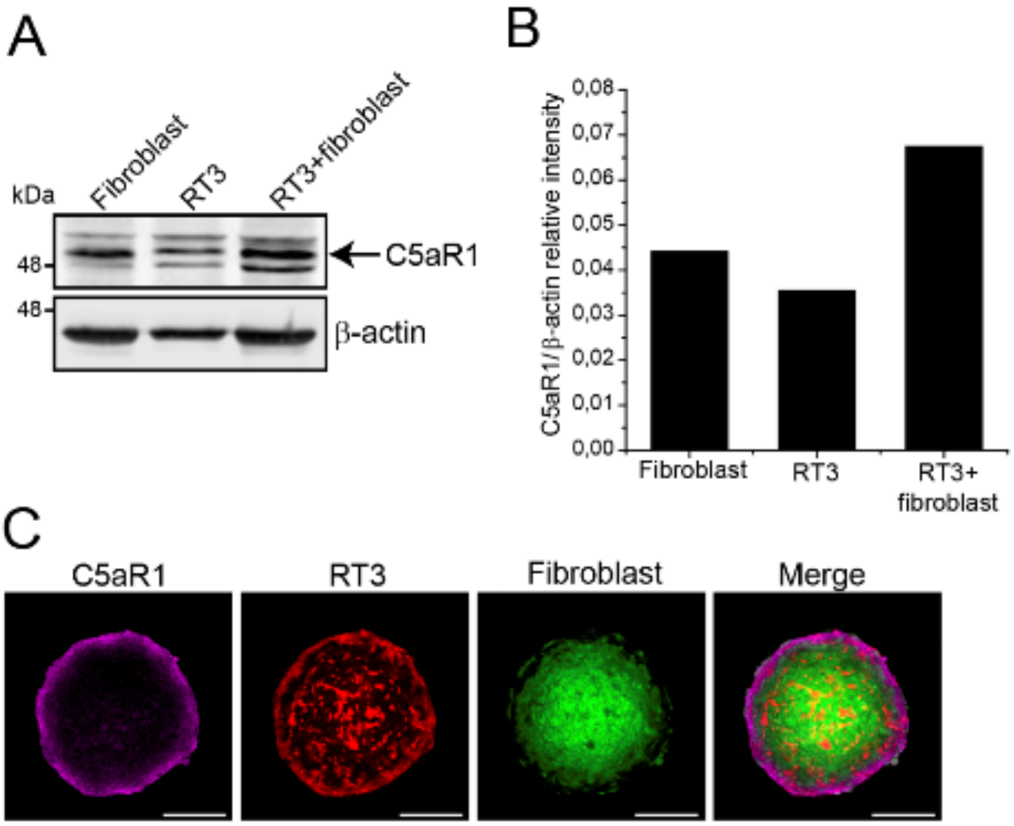

**Figure S1: C5aR1 expression in RT3 cells is induced by co-culture with fibroblasts.**

(A) Western blot analysis of C5aR1 in 3D spheroids composed of human skin fibroblasts, metastastic Ha-*ras*-transformed HaCaT cells (RT3) and their co-coculture. β-actin was used as a loading control. n=2. (B) Quantification of C5aR1 levels from western blots in (A). The graph illustrates C5aR1 relative intensity to β-actin ± SEM. n=2. (C) Confocal images of spheroids with RT3 cells co-cultured with human skin fibroblasts. The cells were initially labeled with CellTrackers (RT3 cells in red and fibroblasts in green) and the spheroids were allowed to grow for three days. Following PFA fixation, the spheroids were subjected to immunofluorescence and stained for C5aR1. Scale bars, 200 µm. n=2, 16 spheroids imaged.

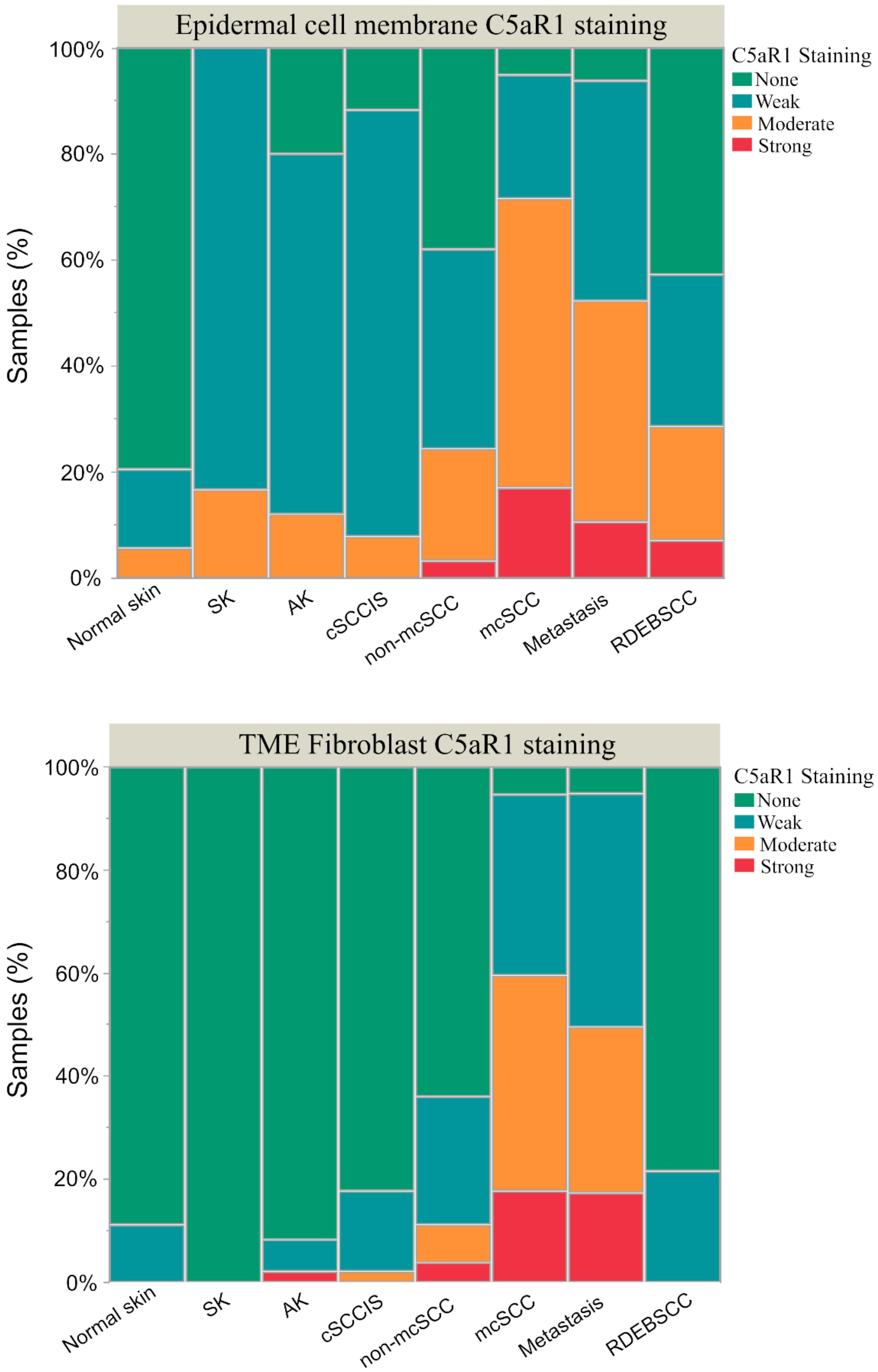

**Figure S2. C5aR1 staining on epithelial cell surface and in TME fibroblasts**.

The original results of semiquantitative analysis of chromogenic IHC samples showing differences of C5aR1 staining on cSCC cell surface and in TME fibroblasts.

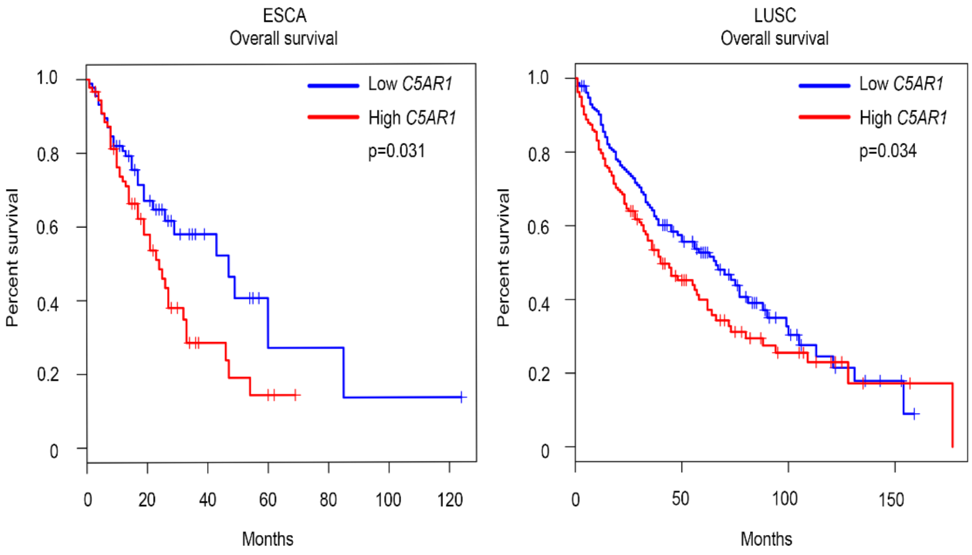

**Figure S3. Correlation of the expression of *C5AR1* with a prognosis of the patients with esophageal carcinoma and lung SCC.**

Evaluation of the impact of *C5AR1* on the overall survival of esophageal carcinoma (ESCA) and lung SCC (LUSC) patients was performed using the GEPIA tool with available TCGA data.

#### Table S1. Baseline characteristics of patients and tumours.

| cSCC patient characteristics | Total<br><br>n=143 patients | non-mcSCC<br><br>n=90 patients | mcSCC<br><br>n=53 patients |
|---|---|---|---|
| **Age (y), mean (min, max)** | 77.3 (27, 102) | 79.3 (47,102) | 74.4 (27, 93) |
| **Gender** | | | |
| **Male, n (%)** | 90 (59.6) | 42 (56.0) | 34 (65.4) |
| **Female, n (%)** | 61 (40.4) | 33 (44.0) | 18 (34.6) |

| cSCC tumor characteristics | Total<br><br>n=152 tumors | non-mcSCC<br><br>n=97 tumors | mcSCC<br><br>n=55 tumors |
|---|---|---|---|
| **Location** | | | |
| **Head and neck, n (%)** | 127 (83.6) | 84 (86.6) | 43 (78.2) |
| **Upper limb, n (%)** | 13 (8.6) | 7 (7.2) | 6 (10.9) |
| **Lower limb, n (%)** | 9 (5.9) | 4 (4.1) | 5 (9.1) |
| **Torso, n (%)** | 3 (2.0) | 2 (2.1) | 1 (1.8) |
| **Diameter (mm), Md (Q1, Q3)** | 15 (9, 30) | 11 (9, 20) | 28 (15, 40) |
| **Differentiation** | | | |
| **Good, n (%)** | 55 (36.2) | 44 (45.4) | 11 (20.0) |
| **Moderate, n (%)** | 65 (42.8) | 36 (37.1) | 29 (52.7) |
| **Poor, n (%)** | 30 (19.7) | 15 (15.5) | 15 (27.3) |
| **Missing, n (%)** | 2 (1.3) | 2 (2.3) | 0 (0) |
| **Necrosis among primary tumors** | | | |
| **No, n (%)** | 134 (88.2) | 90 (92.8) | 44 (80.0) |
| **Yes, n (%)** | 15 (9.8) | 6 (6.2) | 9 (16.4) |
| **Missing, n (%)** | 3 (3.0) | 1 (1.0) | 2 (3.6) |
| **Clark's level** | | | |
| **2-4, n (%)** | 68 (44.8) | 66 (68.0) | 2 (3.6) |
| **5, n (%)** | 80 (52.6) | 29 (29.9) | 51 (92.8) |
| **Missing, n (%)** | 4 (2.6) | 2 (2.1) | 2 (3.6) |
| **Invasion beyond fat** | | | |
| **No, n (%)** | 102 (67.1) | 82 (84.5) | 20 (36.4) |
| **Yes, n (%)** | 49 (32.2) | 15 (15.5) | 34 (61.8) |
| **Missing, n (%)** | 1 (0.7) | 0 (0.0) | 1 (1.8) |
| **AJCC-8** | | | |
| **T1, n (%)** | 58 (42.3) | 58 (70.7) | 0 (0.0) |
| **T2, n (%)** | 6 (4.4) | 6 (7.3) | 0 (0.0) |
| **T3, n (%)** | 22 (16.1) | 15 (18.3) | 7 (12.7) |
| **T4a-T4b, n (%)** | 50 (36.5) | 3 (3.7) | 47 (85.5) |
| **TX, n (%)** | 1 (0.7) | 0 (0.0) | 1 (1.8) |
| **BWH** | | | |
| **T1, n (%)** | 71 (46.7) | 64 (66.0) | 7 (12.7) |
| **T2a, n (%)** | 32 (21.1) | 17 (17.5) | 15 (27.3) |
| **T2b, n (%)** | 39 (25.7) | 13 (13.4) | 26 (47.3) |
| **T3, n (%)** | 9 (5.9) | 3 (3.1) | 6 (10.9) |
| **TX, n (%)** | 1 (0.6) | 0 (0.0) | 1 (1.8) |

**Table S2. Antibodies used in this study.**

| Spheroid Immunofluorescence | Specificity |
|---|---|
| A21071 | Alexa Fluor 633 goat anti-rabbit IgG secondary antibody (A21071, Invitrogen; 1:200 in PBST) |
| Ab234757 | C5aR1 antibody (ab234757, Abcam; 1:100 in PBST) |
| Xenograft IHC | |
| Ab11867 | C5aR1-antibody (C5a-R (S5/1) (Abcam / ab11867) |
| Multiplexed Immunofluorescence | |
| TSA-555 | Ra FAP (ab207178) 1:500 |
| Alexa-647 | Ma C5aR1 (ab11867) 1:100 |
| Alexa-750 | Ma SMA (DAKO M0851) 1:200; O/N 4°C |
| Alexa-647 | R-anti-CD45 (CST13917) 1:100 |
| Alexa-750 | Pan-EPI |
| Chromogenic IHC | |
| Ab11867 | C5aR1-antibody (C5a-R (S5/1) (Abcam / ab11867) |
